# The Silent Threat: Unveiling the Mystery of Post-Op Hypoxemia after Open Heart Surgery

**DOI:** 10.1101/2024.07.25.24311019

**Authors:** Hamdan Mallick, Abdullah Khalid, Yasir Khan, Shiraz Hashmi, Syed Shahabuddin

**Affiliations:** Medical Student, Aga Khan University, Karachi, Pakistan; Section of Cardiothoracic Surgery, Aga Khan University, Pakistan

**Keywords:** Hypoxemia, Cardiac Critical Care, Post-Operative Complications, Predictors, Open Heart Surgery

## Abstract

Hypoxemia is a well-known postoperative complication following open heart surgery. There is minimal literature and a lack of consensus on postoperative hypoxemia as a complication following open heart surgery and its relation to adverse outcomes, risk factors, and population characteristics. This is especially more apparent in lower middle-income countries where there is a double burden of disease. We aimed to determine the predictive and risk factors for postoperative hypoxemia due to the high incidence of associated complications that include increased morbidity, mortality, and length of hospital stay.

This study was a retrospective analysis, evaluating first-time open-heart surgery patients over a period of one year from January to December 2020. Data was collected prospectively through the section of cardiothoracic surgery’s computerized database, using standardized data collection strategies and definitions.

The overall prevalence of hypoxemia was estimated to be 73 (11.8%). Morbidity and Mortality were significantly higher in the hypoxemic group. Male gender, diabetes, hypertension, low EF%, CBP time, increased preoperative PaCO2, were found to be independent risk factors for the development of hypoxemia.

From the results of this study, it could be concluded that the magnitude of hypoxemia is substantial after on-pump open heart surgery. High risk and independent factors identified from this study were cumulatively grouped and preventive use of BiPAP was found to be a possible option to prevent hypoxemia This study highlights the importance of one of the underestimated post op morbid factors that has cost, quality, and outcome implications.

## INTRODUCTION

Open heart surgery is one of the most performed procedures, universally, for a wide variety of coronary artery diseases. In the last decade, numerous advances in anesthesia and surgical techniques such as the prevalent use of membrane oxygenators and improvements in myocardial protection and materials used, have led to earlier extubation and discharge from the hospital [1,2]. However, the systemic inflammatory response syndrome seen in cardiac surgery often leads to organ dysfunction which comprises of complications such as renal failure, acute lung injury and multiple organ dysfunction syndrome [3,4]. The activation of this inflammatory response has shown to cause pulmonary endothelium changes, pulmonary edema, shunt increase and atelectasis, all of which contribute to the manifestation of the postoperative hypoxemia [5], one of the most common postoperative complications, and an important risk factor for reintubation.

Several factors such as old age, increased weight, history of smoking, low ejection fraction, left ventricular dysfunction, and longer cross clamp and pump time have been thought to contribute to the hypoxemia that manifests after a CABG [2,6]. Determining the predictive and risk factors for postoperative hypoxemia is imperative due to the high incidence of associated complications such as; morbidity, increased hospital stay, mortality and cost implication [2]. This is especially important for lower middle-income settings. Very few studies have been done in the region in the subset of the open-heart surgery population. We aimed to determine prevalence, risk factors and short-term outcomes associated with post-open heart surgery hypoxemia. The results of this study will provide a baseline data to help to provide a framework and basis for further follow up studies.

## MATERIALS AND METHODS

This study is a retrospective analysis of prospectively collected data, evaluating first-time open-heart surgery over a period of one year from January to Dec 2020. Data was collected prospectively through the section of cardiothoracic surgery’s computerized database, using a standardized data collection strategy and definitions. A total of 619 patients were included in this study. This analysis excluded patients who underwent redo-cardiac surgery, complex aortic surgeries.

### Study variables

Variables were sub categorized into preoperative, intraoperative, postoperative stages and based on priority of surgery. Baseline characteristics included patient demographics and comorbid conditions such as diabetes mellitus, hypertension, chronic obstructive pulmonary disease (COPD), eGFR, and status of tobacco consumption. Priority of surgery was assessed in the form of elective, urgent, and emergent procedures. Intraoperative variables included cross clamp time, pump time, and type of surgery. For postoperative variables prolonged ventilation was assessed. This study also examined management of hypoxemia and other outcomes in the form of mortality, length of ICU stays, wound infection, length of hospital stay. All given information was collected on a customized proforma.

### Operational definitions

#### Hypoxemia

Postoperative hypoxemia will be assessed through blood oxygen levels by performing arterial blood gas measurements. A level of blood oxygen <70 mmHg on the first postoperative day, within 6 hours after extubation and discharge from CICU.

#### Overweight

Categorized as overweight if BMI ≥ 25. Body-mass index (BMI) will be calculated as weight in kilograms divided by height in meter squared (kg/m2). [NIH]

#### Prolonged Ventilation

Percent of patients aged 18 years and older undergoing isolated CABG who require intubation for more than 24 hours [Society of Thoracic Surgeons]

#### Reopening

Patients who require a return to the operation room for reopening due to any reason.

Risk-Adjusted Surgical Reoperation: Percent of patients aged 18 years and older undergoing isolated CABG who require a return to the operating room for bleeding with or without tamponade, graft occlusion, valve dysfunction, or other cardiac reason [Society of thoracic surgeons]

#### Operative Mortality

Death during the hospital stay or within 30 days following discharge.

### Statistical analysis

Data was entered and analyzed using SPSS version 20. Data was presented as mean with standard deviation; Discrete variables were presented as frequencies and percentages. For numerical data, an independent sample t-test was applied. For categorical data, chi-squared was performed. Stratified analysis was performed by hypoxemia vs. non-hypoxemia groups with pre- and peri-operative variables. Univariate and multivariate logistic regression analysis was performed to identify independent risk factors for postoperative hypoxemia. A p<0.05 is considered as significant.

## RESULTS

### Baseline characteristics

We analyzed 619 patients who underwent a first-time open-heart surgery procedure. Of this given study population, a total of 73 (11.8%) patients developed postoperative hypoxemia.

Baseline characteristics of the population is presented in Table-1. Patients who developed hypoxemia were mostly younger (54.2±16.5, p=0.043) than the non-hypoxemic group (57.3±11.6, p= 0.043). This could be indicated by the finding that most elderly people underwent isolated CABG procedures as opposed to younger patients. Most of the study population was male (77.1/%) and was most likely to develop hypoxemia 47 (64%) as compared to the female 26 (35.6%), p=0.006). In addition, most of the patients in hypoxemia group had a low ejection fraction of <45% as compared to non-hypoxemia group. Preop condition like cardiogenic shock was more in hypoxemic group as compared to non-hypoxemic group, p value for all were <0.001. Pre-Op eGFR was comparable in both the groups however Post Op eGFR was low in hypoxemic group. Comorbid condition like increased eGFR, dyslipidemia and chronic obstructive pulmonary disease, myocardial infarction, history of PCI, and tobacco use were also not significantly different between two groups.

**Table 1:** Baseline characteristics of study population, n=619.

| Characteristics | Hypoxemia |  | P value |
| --- | --- | --- | --- |
| | Yes<br>$n = 73$ (11.8%) | No<br>$n = 546$ (88.2%) | |
| Age in years | $54.2 \pm 16.5$ | $57.3 \pm 11.6$ | 0.043 |
| Male gender | 47 (64.4) | 430 (78.8) | 0.006 |
| Tobacco User | 14 (19.2) | 114 (20.9) | 0.878 |
| Diabetes | 14 (19.2) | 179 (32.8) | 0.022 |
| Hypertension | 20 (27.4) | 245 (44.9) | 0.005 |
| Dyslipidemia | 8 (11.0) | 112 (20.5) | 0.058 |
| COPD | 5 (6.8) | 31 (5.7) | 0.688 |
| Myocardial Infarction | 15 (20.5) | 92 (16.8) | 0.433 |
| History of PCI | 8 (11.0) | 39 (7.1) | 0.248 |
| Ejection Fraction% | $45.3 \pm 9.3$ | $49.39 \pm 8.8$ | $<0.001$ |
| EF ≤ 45% | 56 (76.7) | 114 (20.9) | <0.001 |
| Shock | 7 (9.6) | 5 (0.9) | <0.001 |
| Pre-op eGFR | 66.4 ± 39.5 | 70.3 ± 25.7 | 0.275 |
| Post-op eGFR | 47.2 ± 36.4 | 60.4 ± 28.4 | 0.001 |

COPD: chronic obstructive pulmonary disease, PCI: percutaneous coronary intervention, EF: ejection fraction, GFR: glomerular filtration rate, Pre-op: pre-operative, Post-op: post-operative

### Prevalence of Hypoxemia by type and priority of Procedures: Priority of Procedures

Elective procedures were the most performed procedure in non-hypoxemic group (72.3% vs. 52.1%) followed by urgent (12.1 vs. 38.4%) and emergent (6.6% vs. 9.9%), p=0.002. (Table-2a).

**Table 2a:** Priority of Procedures, n=619.

| Priority of Surgery | Hypoxemia |  | P value |
| --- | --- | --- | --- |
|  | Yes<br>n=73 (11.8%) | No<br>n=546 (88.2%) |  |
| Elective | 38 (52.1) | 395 (72.3) | 0.002 |
| Urgent | 28 (38.4) | 115 (21.1) |  |
| Emergent | 7 (9.6) | 36 (6.6) |  |

Isolated CABG was the most performed procedure overall. However, the proportion of procedures performed in hypoxemic group was lower in the hypoxemic group versus the non-hypoxemic group (57.5% vs. 75.5%). This was followed by comparable numbers for isolated valve replacements (13.7% vs 13.6%) in both the groups. Combined CABG and valve replacements (6.8% vs 2.4%), double valve replacement (2.7% vs 1.3%) and other cardiac procedures (19.2% vs 5.3%) such as aneurysm repair, congenital heart defect repairs were found to be more performed in the hypoxemic group, overall p<0.001. (Table 2b)

**Table 2b:** Type of Procedures performed, n=619.

| **Type of Procedures** | **Hypoxemia** |  | **P value** |
| --- | --- | --- | --- |
|  | Yes<br>n=73 (11.8%) | No<br>n=546 (88.2%) |  |
| Isolated CABG | 42 (57.5) | 423 (77.5) | <0.001 |
| Isolated Valve replacement | 10 (13.7) | 74 (13.6) |  |
| CABG and valve replacement | 5 (6.8) | 13 (2.4) |  |
| Double Valve Replacement | 2 (2.7) | 7 (1.3) |  |
| Other cardiac procedures | 14 (19.2) | 29 (5.3) |  |

CABG: Coronary Artery Bypass Grafting

Interestingly all surgeries that were not an isolated CABG were associated with a higher incidence of hypoxemia (p<0.001). Prevalence of postoperative hypoxemia was greater in patients who underwent urgent (19.6%, p<0.002) and emergent (16.27%, p<0.002) cases as compared to those patients who underwent elective cases (8.78%, p<0.002) (Table 2a-2b).

### Perioperative findings and Management

Intra operative included the mean pump and cross clamp time (Table-3a). The mean pump time (minutes) was higher in hypoxemic patients 125.1 ±40.0, compared to non-hypoxemic patients 98.01 ± 26.5, p<0.001). Cross clamp time was also found to be greater in hypoxemic patients 80 ± 33.3, than in those patients who did not develop hypoxemia 73.63 ± 22.4, p=0.034 (Table-3a).

**Table 3a:** Intraoperative variables, n=619.

| Management | Hypoxemia |  | P value |
| --- | --- | --- | --- |
|  | Yes<br>n=73<br>(11.8%) | No<br>n=546 (88.2%) |  |
| Pump time | $125.1 \pm 40$ | $98.01 \pm 26.5$ | $<0.001$ |
| Cross clamp time | $80 \pm 33.3$ | $73.63 \pm 22.4$ | 0.034 |

**Table 3b:** Postoperative findings, n=619.

| Management | Hypoxemia |  | P value |
| --- | --- | --- | --- |
|  | Yes<br>n=73 (11.8%) | No<br>n=546 (88.2%) |  |
| Prolonged ventilation | 12 (16.4) | 16 (2.9) | $<0.001$ |
| PCO<sub>2</sub> mmHg | $54.3 \pm 13.8$ | $45.91 \pm 5.9$ | $<0.001$ |
| PO<sub>2</sub> mmHg | $44 \pm 11.6$ | $110.26 \pm 40.3$ | $<0.001$ |

PCO<sub>2</sub>: Partial pressure of carbon dioxide, PO<sub>2</sub>: Partial pressure of oxygen.

A considerable proportion of Hypoxemic patients required Prolonged ventilation (16.4% vs., 2.9%), p <0. 001. Mean PO2 mmHg was low (44 ± 11.6 vs. 110.26 ± 40.3) and PCO2 mm Hg (54.3 ± 13.8 vs. 45.91 ± 5.9) was high in hypoxemic population, p<0.001 for both. Post-operative GFR was less in the hypoxemic group (47.2+36.4, P<0.001) compared to the non-hypoxemic group (60.4+28.4, p<0.001). (Table-3)

Management was conducted through 3 methods: BIPAPS, Reintubation, and non-invasive post-op management. BIPAP was the most common management option used for the management of hypoxemia. Of these, 38 (52.1%) were in hypoxemic group and 91 (16.7%) were in non-hypoxemic group, followed by simple post-op management 29 (39.7% vs. 82%), and finally re-intubation 6 (8.2% vs. 0.9%), p value for all <0.001 (Table-4). These management strategies were based on the overall postoperative condition of the patient, vitals, and the physician’s discretion.

**Table 4:**
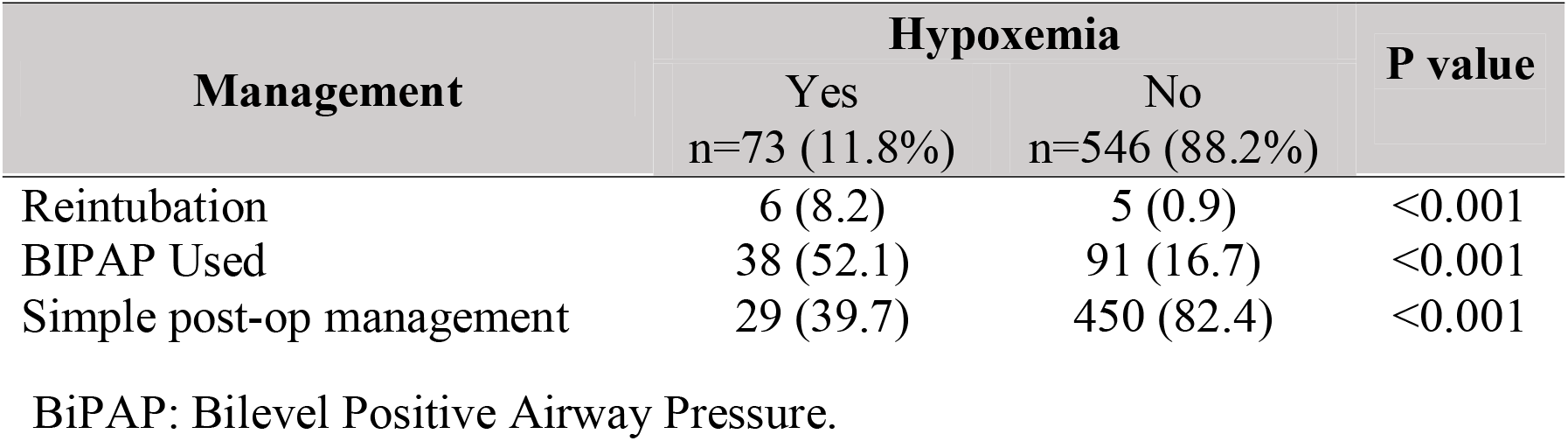
Post-op management, n=619.

### Outcome Measures

Hypoxemic patients were associated with a greater likelihood of overall morbidity than non-hypoxemic patients (P<0.001). The rates of mortality were also significantly greater in hypoxemic patients as compared to non-hypoxemic patients p<0.001. Group difference on Prolonged ICU stay (hrs.), rates of deep sternal wound, a-fib, reopening of patient, and cerebrovascular complications could not be determined in this subset of population. p>0.05.

Hypoxemic patients were more likely to get readmitted (9.6%) as compared to the counterpart (3.8%), p=0.027. Length of hospital stay in hypoxemic patients was 12.9 ± 7.6 compared to non-hypoxemic group and 8.4 ± 7.6, p<0.001). Readmission was required in 30 patients, (9.6% vs. 3.8%), p=0.027. Overall morbidity was seen in 211 patients, (56.2%) vs.31.1%) p<0.001. There were a total 18 operative deaths, 12(16.4%) vs. 6 (1,1) in both groups respectively. (Table-5).

**Table 5:** Peri-Operative variables and outcomes, n=619.

| Peri-Op variables | Hypoxemia |  | P value |
| --- | --- | --- | --- |
|  | Yes<br>n=73 (11.8%) | No<br>n=546 (88.2%) |  |
| CIUC stay (hrs) | $52.1 \pm 18.7$ | $54 \pm 28.5$ | 0.572 |
| Length of hospital stay | 12.9 ± 7.6 | 8.4 ± 7.6 | <0.001 |
| Overall morbidity | 41 (56.2) | 170 (31.1) | <0.001 |
| Prolonged ventilation | 12 (16.4) | 16 (2.9) | <0.001 |
| Atrial Fibrillation | 13 (17.8) | 90 (16.5) | 0.775 |
| Reopen due to any reason | 1 (1.4) | 5 (0.9) | 0.710 |
| Prolonged ICU stay (hrs) | 57.1 ± 22.4 | 50.6 ± 35.2 | 0.233 |
| Deep Sternal wound | 3 (4.1) | 7 (1.3) | 0.072 |
| Cerebrovascular events | 3 (4.1) | 7 (1.3) | 0.072 |
| Readmission | 7 (9.6) | 21 (3.8) | 0.027 |
| In hospital Mortality | 12 (16.4) | 6 (1.1) | <0.001 |

CICU: Cardiac Intensive Care Unit, ICU: Intensive Care Unit

### Predictors of Hypoxemia

After applying multiple regression analysis, the following were found to be independent risk factors for developing postoperative hypoxemia :preoperative diabetes (odds ratio (OR)=2.52, 95% confidence interval (CI)1.01– 6.3),hypertension (OR=2.62, 95% CI 1.2–5.76), EF% (OR=0.94, 95%CI 0.91– 0.98), Age (OR= 1.00, 95% CI 0.98–1.02), Gender (OR=1.48, 95%CI 0.77– 2.86), CBP time (OR=1.03, 95%CI 1.02–1.04), preoperative PaCO2 (OR= 1.07, 95% CI 1.03–1.1), length of hospital stay (OR=1.113, 95%CI 1.07–1.19) after adjusting for all variables in the model. (Table-5).

## DISCUSSION

First 24 postoperative hours are critical for major surgeries. It is found in our setup that most patient becomes hypoxemic within 24 hours, hence requiring noninvasive ventilation. However, data is scarce on the prevalence and incidence of hypoxemia after Open heart Surgery. In the loco-regional context no study is found in the literature and most of the literature is from western countries. The cardiovascular diseases (CVD) risk profile of their population varies from our population. In south Asia the conventional CVD risks are more prevalent [7]. Therefore, this study is designed to determine the frequency of hypoxemia in patients undergoing Open heart Surgery after extubation on 1st postoperative day. Hypoxemia is defined as a condition where the arterial oxygen tension or partial pressure of oxygen (PaO2) falls below the normal range [8]. The PaO2/FiO2 ratio is one of the most important indicators in assessing the severity of pulmonary injury of mechanically ventilated patient. The findings of this study will help to formulate strategies to minimize the complication which eventually will decrease postoperative hospital stay, cost of care and surgical outcomes. Throughout this study, the effects of acute hypoxemia on short-term prognosis including ICU length of stay, intubation duration, morbidity, and mortality was studied. Due to an inadequacy of facilities, the ABGs were the only criterion for including patients in the hypoxemia class. It was reported that the average of PaO2 for hypoxemic patients was 44 ± 11.6 mm Hg. Based on the PaO2 values, only 11.8% of the patients developed postoperative hypoxemia, whereas the rest had normal respiratory conditions. Along with the observed increase in ventilation time and length of hospital time in these patients which was expected based on a study by Weiss et al. [2], statistical differences in morbidity and mortality were also observed. This is the main result of this study. The percentage of patients who developed hypoxemia (11.8%) is much less than the data reported previously, which ranged from 18.8-27.08% [9].One outlier and stark contrast was the 55% hypoxemia reported by dos Santos et al. [6]. This may be attributed to the significant overweight population and presence of obesity, diabetes, and hypertension as significant comorbidities. Though diabetes and hypertension were found to be independent risk factors for the development of postoperative hypoxemia, the low prevalence of these comorbidities may have contributed to better postoperative outcomes. Following our univariate analysis, the following risk factors were determined: Male gender, Low LVEF, cardiogenic shock, mean pump, cross clamp time, emergent & urgent surgery, and all surgeries besides simple isolated CABG. Additionally, through logistic regression, the following independent risk factors of postoperative hypoxemia were found; diabetes and hypertension, recent MI, low EF% and CPB time.

Low ejection fraction was an independent predictive factor identified by our study; this is backed up by a previous study done by Szeles TF et al. The extent of impact of the Left ventricular dysfunction on increasing the risk of severe hypoxemia was almost doubled (OR=1.835) [2]. When comparing the impact of EF% in the development of hypoxemia following logistic regression analysis in this study, the association was less profound and an odds ratio of 0.96.

CPB time was another independent risk factor that was found in our study with an odds ratio of 1.02. The study by Szeles TF et al. interestingly separated the CPB time factor into 3 groups fulfilled by either CPB>120 min, CPB<120 min, and without CPB. CPB>120 min and CPB<120 min was both found to be significant risk factors following multivariate analysis. In fact, the odds ratio increased from 2.31 to 3.195 when comparing the CPB<120 min to CPB>120 minutes [2]. This was a clear indication of the significant role as an indicator for the development of postoperative hypoxemia that is also found following our analysis. Though we may not be able to compare the raw odd ratio due to the different criteria for the CBP groups. There is a clear indication and support for this claim. A study by Qiang Ji et al. also reported an association through univariate analysis and reported CPB time as a risk factor [10]. The mean CPB time for both hypoxemic and non-hypoxemic groups however were both higher (116.60±22.96, 97.86±14.69) compared to our study (80±33.3, 73.63±22.4). This may be due to the advances in CPB techniques and superior equipment that was found in 2008.

History of an acute myocardial infarction was an independent risk factor in this study with an OR of 1.28. The study by Qiang Ji et al. showed a much stronger correlation with an odds ratio of 3.351 [10].

Diabetes was determined to be an independent risk factor for the development of hypoxemia. This is a phenomenon that can be explained by the discovery that the lungs are one of “target organs” of diabetes [11]. The pathophysiology due to hyperglycemia may be attributed to an amalgamation of contributing factors. A structural change develops as the endothelial permeability increases, leading to leakage of micromolecules proteins and plasma. This is further worsened by the glycosylation of tissue protein and accrementition of connective tissue which decreases the overall elastance of the lung. The additional impairment and disruption of vascular epithelium leads to thickening of the pulmonary basement membrane and an increase in diffusion coefficient. These factors result in a decreased ventilation rate and diffusion capacity leading to an overall decrease in lung functionality [10]. Another significant comorbid that was found to be an independent risk factor: hypertension. The existing literature has not made a significant association between hypertension and the development of postoperative hypoxemia. Hypertension has multiple effects on PFTs and normal respiration. A study Margretardottir et al. reported that hypertensives had a greater propensity to have a persistently lower FEV1% and FVC% [9].

This decrease in pulmonary function may be described as the direct result of the increased pulmonary and systemic vascular resistance leading to an increased vascular thickness. This is accompanied by the loss of elasticity in the pulmonary vascular tree, irrespective of any parenchymal changes in the pulmonary system. This pathological change harms normal physiological function and leads to a decreased vital capacity and FEV1 [12].

Smoking, a risk factor that has been heavily reported in previous studies [6,13], was not a significant contributor to the development of postoperative hypoxemia in our study, perhaps due to the difference in age groups in our sample size.

COPD is a significant risk factor for higher mortality in general and may be part of a chronic systemic inflammatory response [9]. This was supported and found to be an independent risk factor for the development of hypoxemia postoperatively by Qiang Ji et al. [10]. Our study, however, did not show comparable results and COPD was not an independent risk factor. In our study, this is due to use of BiPAP immediately after exhumation in that subset of population. This immediate use of BiPAP will also decrease the incidence of re-intubation [14]. Prolonged ICU stay was not seen in our patient groups. These results coincide with other recent studies [5,6,13]. On the existing studies pertaining to this topic, management has not been discussed in literature thoroughly. Our management scope attempted to deal with patients through reintubation, BiPAP, and simple post-operative management. We found that 52.1% of the patients who developed postoperative hypoxemia required use of BIPAP. This was in comparison to the 16.7 % that was done in the non-hypoxemic group. The aggressive noninvasive approach of using iIPAP in our smoker, COPD, and asthma patients immediately after re-intubation early and reintubation may have been the reason for our decreased morbidity and mortality rates; Since, these interventions were not used in our diabetic, hypertensive, and heart failure patients. This use of BiPAP could be a preventive strategy in the future for the prevention of hypoxemia [15]. Hypoxemia was a significant predictor for in hospital mortality, as up to 16% of those patients passed away due to chronic respiratory distress syndrome. This is in stark contrast to studies where death was not an exclusion criterion the mortality rate is minimal or near 0% [1,5,16].

### Strengths and Limitations

To our knowledge this is the first study that estimates the prevalence and predictors in open heart surgery including all types of procedures. The data was prospectively collected on a large sample though analysis was retrospective. We used the standard definition recommended by the Society of Thoracic Surgery for the outcomes. However, the study was a single center estimation of hypoxemia and could not be generalizable to other populations in the region with different risk profiles and preferences of management. In addition, we had decided not to investigate preexisting causes of hypoxemia prior to admission, as the study was designed for an inpatient setting.

Another limitation with regards to the hospital stay of the patients was that their cost of stay was not considered, which could have indicated their socioeconomic background.

## CONCLUSION

From the result of this study, it could be concluded that the magnitude of hypoxemia is substantial after on-pump open heart Surgery. procedures. Male gender, diabetes, hypertension, low EF%, CBP time, increased preoperative PaCO2, were found to be independent risk factors for the development of hypoxemia. High risk and independent factors identified from this study were cumulatively grouped and preventive use of BiPAP was found to be a possible option to prevent hypoxemia. This study highlights the importance of one of the underestimated post op morbid factors that has cost, quality, and outcome implications.

## Data Availability

All data produced in the present work are contained in the manuscript

## Ethics approval and consent to participate

The study was approved by the Institutional Review Board at Aga Khan University.

## Availability of data and materials

Data can be provided upon reasonable request.

## Acknowledgements

None

